# Human metabolic chambers reveal a coordinated metabolic-physiologic response to nutrition

**DOI:** 10.1101/2024.04.08.24305087

**Authors:** Andrew S. Perry, Paolo Piaggi, Shi Huang, Matthew Nayor, Jane Freedman, Kari North, Jennifer Below, Clary Clish, Venkatesh L. Murthy, Jonathan Krakoff, Ravi V. Shah

## Abstract

The emerging field of precision nutrition is based on the notion that inter-individual responses across diets of different calorie-macronutrient content may contribute to inter-individual differences in metabolism, adiposity, and weight gain. Free-living diet studies have been traditionally challenged by difficulties in controlling adherence to prescribed calories and macronutrient content and rarely allow a period of metabolic stability prior to metabolic measures (to minimize influences of weight changes). In this context, key physiologic measures central to precision nutrition responses may be most precisely quantified via whole room indirect calorimetry over 24-h, in which precise control of activity and nutrition can be achieved. In addition, these studies represent unique “N of 1” human crossover metabolic-physiologic experiments during which specific molecular pathways central to nutrient metabolism may be discerned. Here, we quantified 263 circulating metabolites during a ≈40-day inpatient admission in which up to 94 participants underwent seven monitored 24-h nutritional interventions of differing macronutrient composition in a whole-room indirect calorimeter to capture precision metabolic responses. Broadly, we observed heterogenous responses in metabolites across dietary chambers, with the exception of carnitines which tracked with 24-h respiratory quotient. We identified excursions in shared metabolic species (e.g., carnitines, glycerophospholipids, amino acids) that mapped onto gold-standard calorimetric measures of substrate oxidation preference and lipid availability. These findings support a coordinated metabolic-physiologic response to nutrition, highlighting the relevance of these controlled settings to uncover biological pathways of energy utilization during precision nutrition studies.

## Introduction

The emerging field of precision nutrition is based on the notion that inter-individual responses across diets of different calorie-macronutrient content may contribute to inter-individual differences in metabolism, adiposity, and weight gain. Studies from our group and others have focused on two major metabolic physiologies—variability in fuel selection (substrate oxidation) and diet-induced thermogenesis (energy expenditure, EE) during consumption—as susceptibility phenotypes in weight regulation^1–3^. Many human studies have employed prolonged fasting, caloric restriction^1, 4–6^, or shorter versus longer term responses to macronutrient feeding^7–11^ to start to discern potential contributors to these metabolic phenotypes (e.g., human genetics^12, 13^, resting metabolic rate^1^, substrate oxidation, thermogenesis^14^). Nevertheless, free-living diet studies have been traditionally challenged by difficulties in controlling adherence to prescribed calories and macronutrient content and rarely allow a period of metabolic stability prior to metabolic measures (to minimize influences of weight changes). In this context, key physiologic measures central to precision nutrition responses may be most precisely quantified via whole room indirect calorimetry over 24-h, in which precise control of activity and nutrition can be achieved. In addition, these studies represent unique “N of 1” human crossover metabolic-physiologic experiments during which specific molecular pathways central to nutrient metabolism may be discerned. Here, we quantified 263 circulating metabolites during a ≈40-day inpatient admission in which up to 94 participants underwent seven monitored 24-h nutritional interventions of differing macronutrient composition in a whole-room indirect calorimeter to capture precision metabolic responses. Broadly, we observed heterogenous responses in metabolites across dietary chambers, with the exception of carnitines which tracked with 24-h respiratory quotient. We identified excursions in shared metabolic species (e.g., carnitines, glycerophospholipids, amino acids) that mapped onto gold-standard calorimetric measures of substrate oxidation preference and lipid availability. These findings support a coordinated metabolic-physiologic response to nutrition, highlighting the relevance of these controlled settings to uncover biological pathways of energy utilization during precision nutrition studies.

## Results

Figure 1 shows our experimental design. Nutritional interventions (with macronutrient composition in Figure 1 and **Supplemental Table 1**) included an “energy balance” chamber (to determine an individual’s caloric needs at balanced intake = expenditure), fasting, and serial diets with differing macronutrient composition with 200% caloric excess relative to energy balance. The 200% caloric excess was used as a physiologic probe to elicit phenotypes defined by 24-h EE and 24-h substrate oxidation rate^14^. Dietary intervention chambers after energy balance were performed in random order with an intervening 3 day wash-out period. We used indirect calorimetry during chambers to quantify 24-h EE and substrate oxidation preferences (24-h respiratory quotient [RQ], lipid, carbohydrate, and protein oxidation rate), as described^14^. This design allowed efficient, precise control over confounding (e.g., activity level, sleep, diurnal effects, weight) to quantify phenotypes that capture interindividual heterogeneity in metabolism.

**Figure 1.**
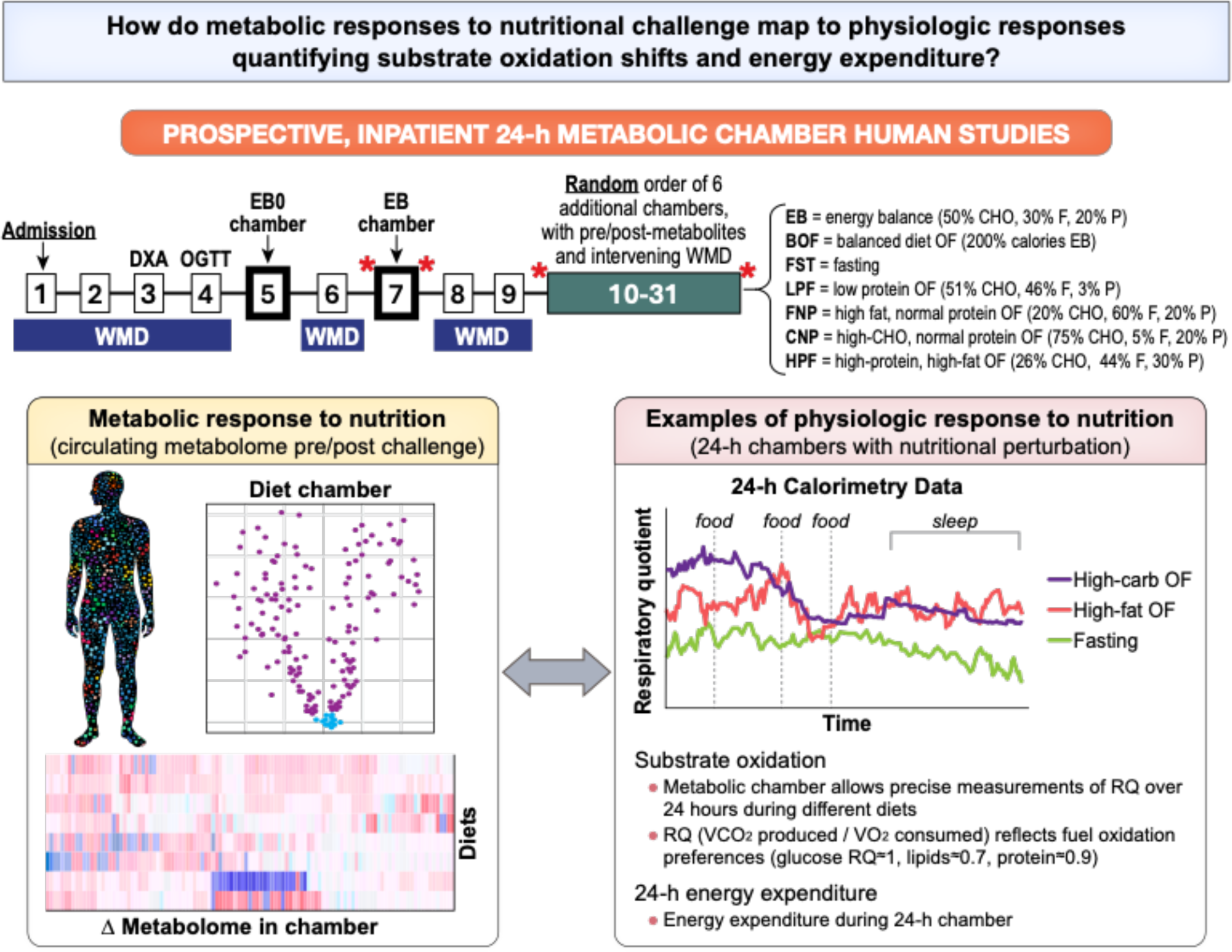
Study scheme, detailing inpatient clinical research protocol and study aims.

The characteristics of our study group are shown in **Supplemental Table 2**. Overall, our population was middle aged (median age 38, 20% female), with a mildly elevated body mass index (median BMI 26 kg/m^2^). Of the 96 participants in our study, 49 participants completed all 7 dietary chambers, and 28 completed 6 chambers (**Extended Data Figure 1**). High carbohydrate overfeeding induced greatest increase in 24-h EE. Substrate oxidation rate changes were generally consistent with the availability of macronutrients (Figure 2): for example, we observed a shift toward fat oxidation (in lieu of carbohydrate) as quantified by lipid oxidation rates and a fall in 24-h RQ in fasting and high-fat diets (with the converse in diets with greater carbohydrate content).

**Figure 2.**
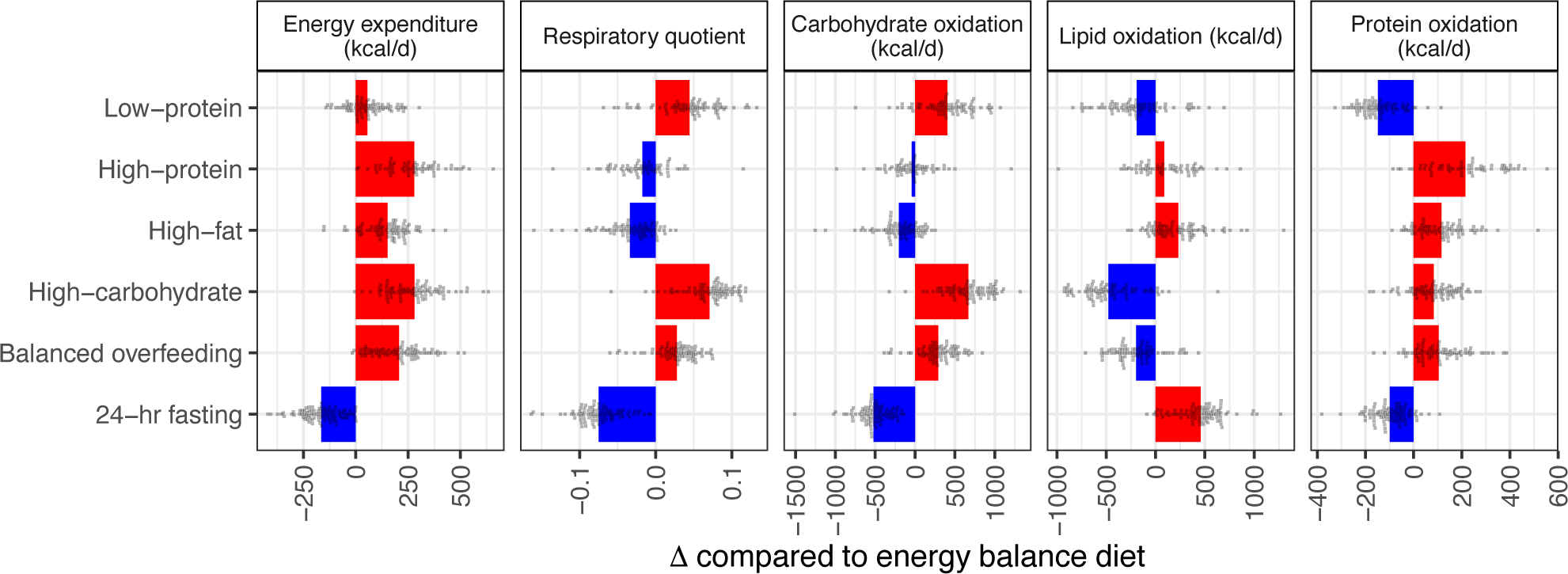
Heterogeneity in metabolic responses to dietary perturbation. Here, we present changes in 5 metabolic parameters (rows) from all dietary chambers, with comparison to the energy balance chamber. Bars represent the mean change across all subjects, and points represent individual participants. While most participants followed the average trend, some individuals displayed opposing changes (e.g., in the high-fat chamber, the average change in respiratory quotient was a decrease, however, in some participants the respiratory quotient increased). Red indicates the average effect was an increase in the metabolic parameter during the dietary chamber compared to energy balance, blue indicates a decrease.

We identified substantial shifts in the assayed circulating metabolome across different 24-h chambers, broadly consistent with putative effects of macronutrient composition in each dietary prescription (Figure 3A-B; full results in **Extended Data Figure 3, Supplemental Table 4**). The 24-h fasting condition elicited the most distinct metabolic pattern consistent with known physiology of early starvation, including increases in mitochondrial β-oxidation (increased acylcarnitines, pantothenic acid), increased ketogenesis (e.g., C4:0-OH carnitine, a ketone body^10^), and increased fatty acid availability (e.g., catabolism of phospholipids [PC/PEs] to diacylglycerols and sphingomyelins^15, 16^). Unexpectedly, we observed a global decrease—not increase—in lysophosphatidylcholines (LPCs) and lysophosphatidylethanolamines (LPEs) during a 24-h fast (another byproduct of PC catabolism), suggesting a complex pathway of phospholipid metabolism during starvation. Indeed, increased LPCs have been linked to decreased fatty acid oxidation^17^ and broad pro-inflammatory phenotypes^18^. Finally, we observed a decrease in many glucogenic amino acids, consistent with increased liver gluconeogenesis^19^ or decreased intake, except for branched-chain amino acids (leucine, isoleucine, valine; BCAAs). These increases in circulating BCAA are consistent with inhibition of their catabolism via increased fatty acid oxidation (as noted above, via NADH availability) and decreased availability of reaction substrates for BCAA catabolism provided through glycolytic flux (not measured here)^19^.

**Figure 3.**
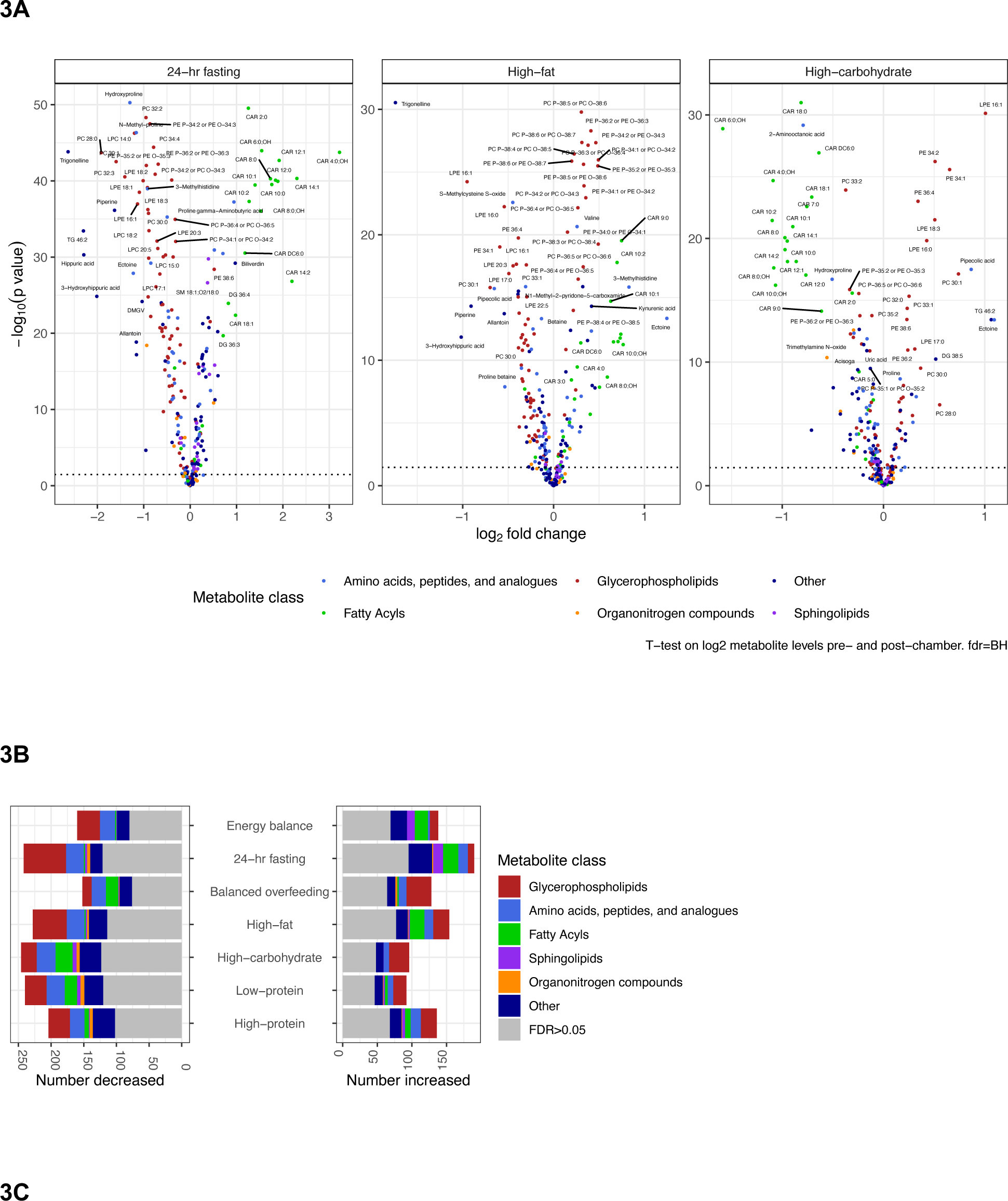
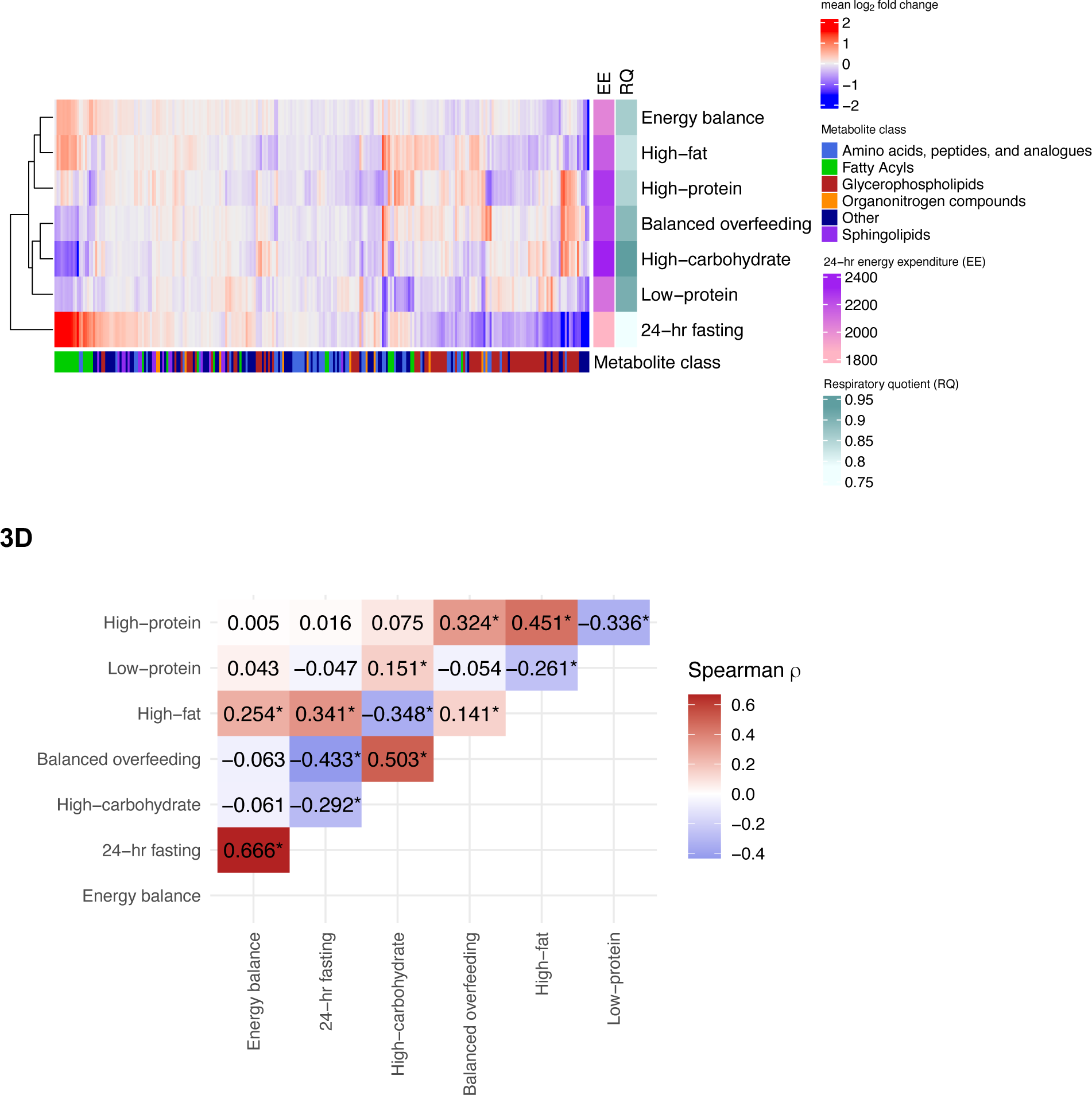
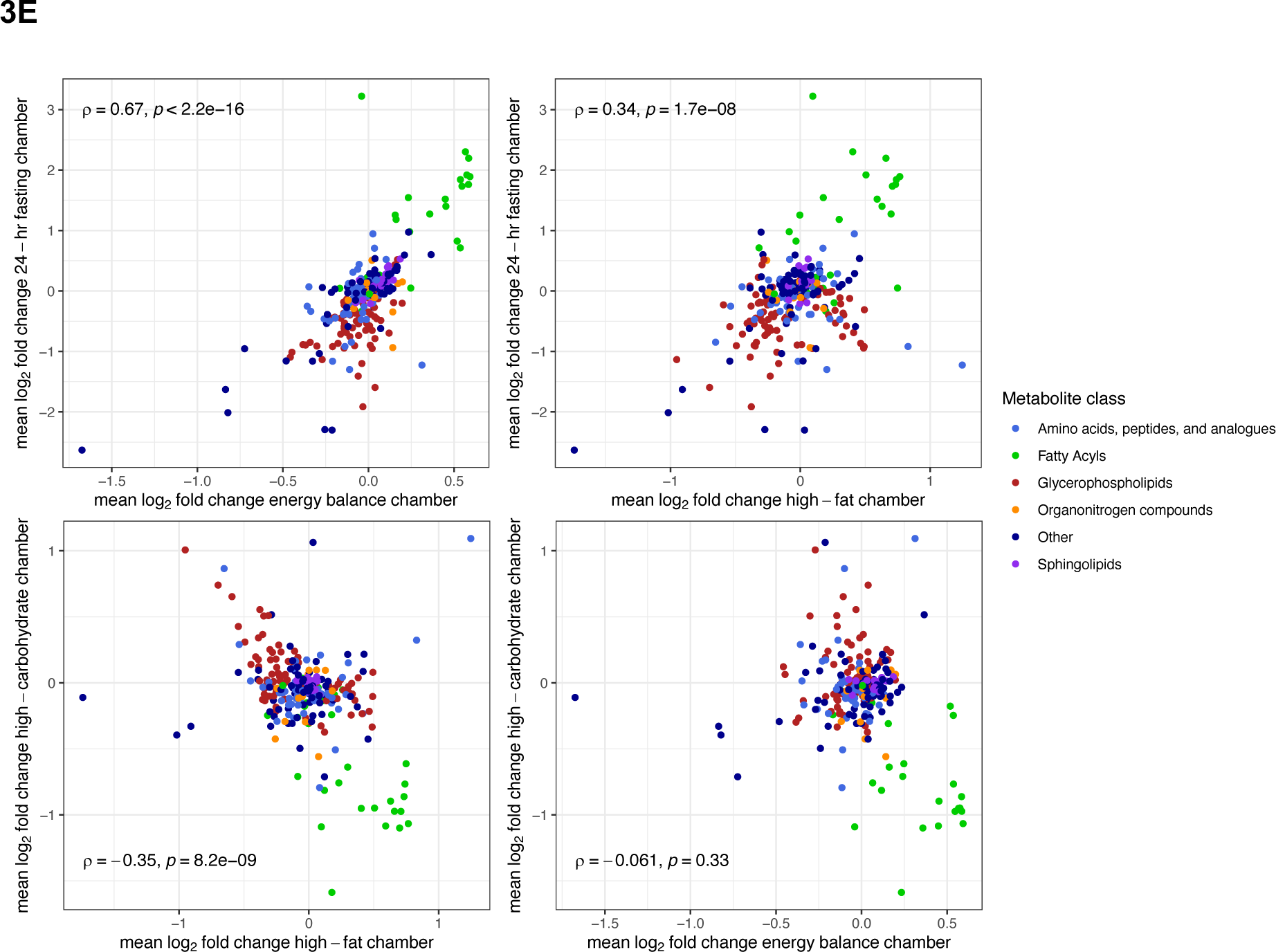
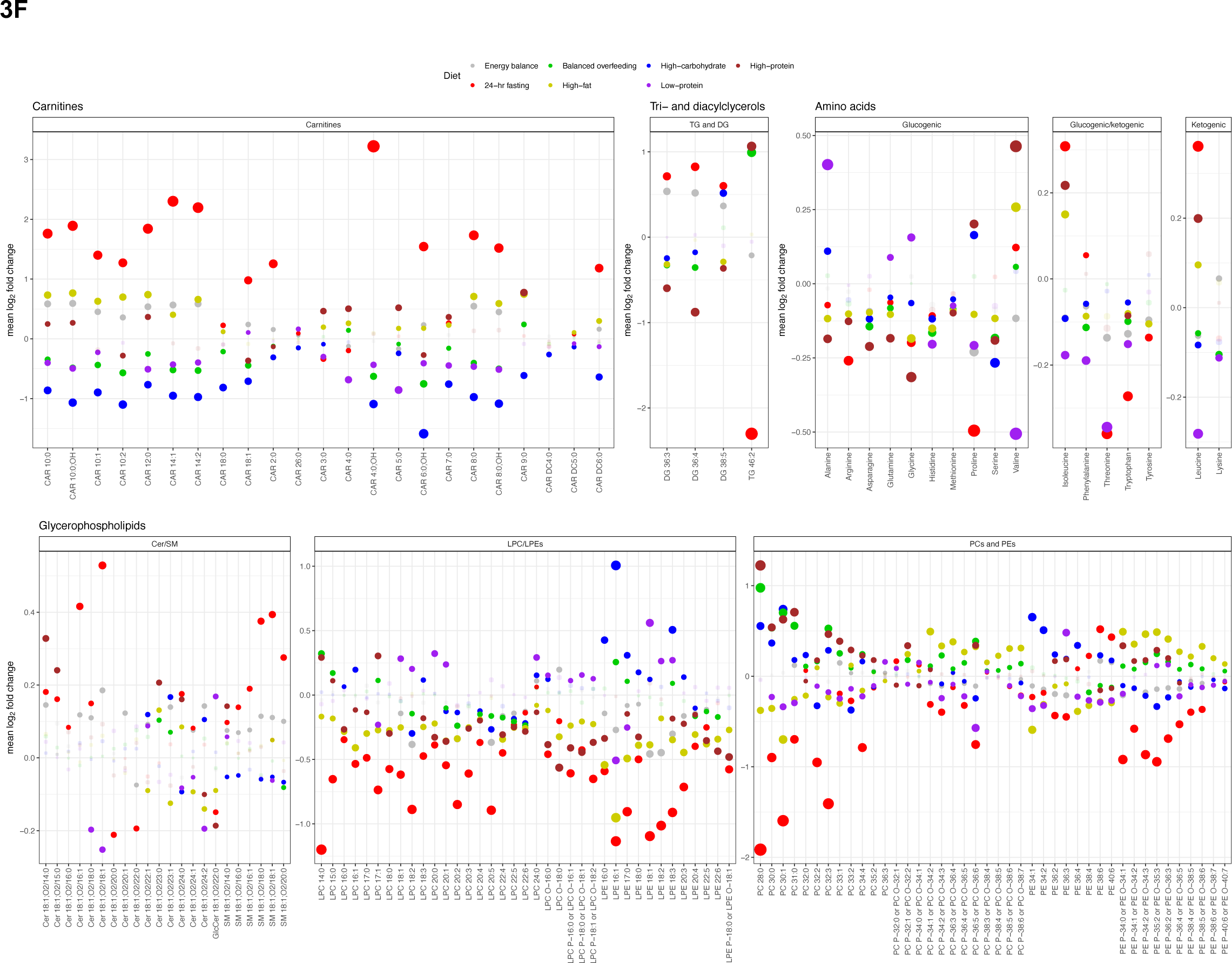
Metabolic responses to dietary intervention and their relation across dietary exposure. **(A)** Volcano plots of paired *t*-tests comparing post-chamber metabolite levels with pre-chamber metabolite levels for select chambers. Results for all chambers are presented in **Extended Data Figure 2**. **(B)** Results from paired *t*-tests are summarized for each dietary chamber, grouped by HMDB metabolite class demonstrating glycerophospholipids as the most common class of metabolite to change. **(C)** Heatmap of the mean log_2_-fold change for all metabolites for all dietary chambers, demonstrating similarities between clusters of diets. **(D)** Spearman correlation heatmap of the paired *t*-test results across dietary chambers. As might be expected, we observed a moderate degree of correlation between the high-carbohydrate and balanced overfeeding diets. **(E)** Select examples from **Panel C** demonstrating correlation of log_2_-fold changes between diets. **(F)** Comparison of the mean log_2_ fold change of metabolites across all 7 dietary chambers. Colors represent the dietary chamber, size is proportional to mean log_2_-fold change, and points are faded if the mean log_2_-fold change for each diet-metabolite combination did not reach at an FDR<5%.

Across all dietary chambers, we observed a generally diverse pattern of metabolite changes, most prominently in amino acids, glycerophospholipids, and fatty acids (Figure 3B). Of note, there was consistency in dietary metabolite responses between chambers that mapped onto 24-h respiratory quotient, a measure of substrate preferences during the chamber (Figure 3C): diets associated with a generally lower 24-h RQ (favoring more fat oxidation) exhibited increases in fatty acids (specifically acylcarnitine species) and decreases in glycerophospholipids (predominantly PCs and their precursor Pes^20^ or catabolic byproducts LPCs/LPEs). Patterns of metabolite excursions (not necessarily overall fold change magnitude) were similar across several diets with closer macronutrient composition, energy intake, or substrate oxidation preferences: metabolic patterns during fasting were correlated directly to energy balance (a lower caloric intake than other chambers, ρ=0.67) and high fat (similar preference for lipid oxidation, ρ=0.34). On the other hand, the relation in metabolic patterns between fasting and diets with greater carbohydrate availability were inverse (high-carbohydrate, ρ=-0.29; balanced overfeeding, ρ=-0.43; Figure 3D-E; (full correlation plots in **Extended Data Figure 3**). Moreover, high protein and high fat diets exhibited concordance, likely owing to a similar fat macronutrient composition (20% vs. 30%, respectively). Given the random order, cross-over design of chambers after energy balance across the study, the consistent mapping of metabolic responses to physiologic adaptation phenotypes and consistent results in mixed modeling results (accounting for carry-over design, **Extended Data Figure 4, Supplemental Table 5**) increased our confidence in absence of a major bias by carry-forward effects of metabolites across chambers. Of note, the low protein chamber exhibited the least concordance in pattern across chambers (Figure 3C). Metabolic changes within the low protein chamber included a generalized decrease in most essential amino acids (see **Extended Data Figure 5**), except alanine, glutamine, and glycine, some of which was consistent with free-living studies of low protein diet^21^. In addition to metabolites that denote substrate utilization, we observed dynamicity in several metabolites of more general clinical interest, specifically microbial products (e.g., trimethylamine N-oxide, indole derivatives, hippuric acid), highlighting unique interconnections among macronutrient intake, host commensal processing and metabolic response^22^. Overall, the metabolite-physiologic adaptation concordance was observed across multiple macronutrient compositions, reinforcing links between individual substrate preferences and metabolic responses to diet.

Given these results suggesting metabolic changes at 24-h of overfeeding diets mirror physiologic substrate oxidation preferences (by 24-h RQ), we explored this directly across diets (full results in **Supplemental Table 6**). Using mixed models across all participants and chambers to maximize power, we identified metabolites associated with 24-h RQ, individual measures of substrate oxidation rates and 24-h EE (Figure 4A). This analysis was predicated on evidence indicating that—although fuel preference is driven primarily by dietary macronutrient content—there remains an intrinsic <u>intra</u>-individual predisposition to fuel preference across diets^14, 23^. Similar to metabolic excursions across diets, <u>changes</u> in carnitines (decreased 24-h RQ) and glycerophospholipids (e.g., PCs and LPCs; increased 24-hr RQ) were associated with substrate preferences, consistent with acylcarnitines as indicators of increased β-oxidation and with a role for PCs as a fatty acid source^24^. These results were robust to regression within each diet separately (**Extended Data Figure 6, Supplemental Table 7**). In addition, we observed a strong, concordant, synergistic relation between the pre-chamber metabolite level and metabolite change during a chamber with the 24-h RQ (ρ=0.86; **Extended Data Figure 7**), driven by “pre-meal” metabolites reflecting fatty acid oxidation (carnitines, pantothenic acid) and lipid availability (PC, LPC/LPE). Pending larger study validation, these results suggest a potential for “resting” pre-meal metabolomics to inform substrate preference which may allow future phenotyping without such detailed interventions.

**Figure 4.**
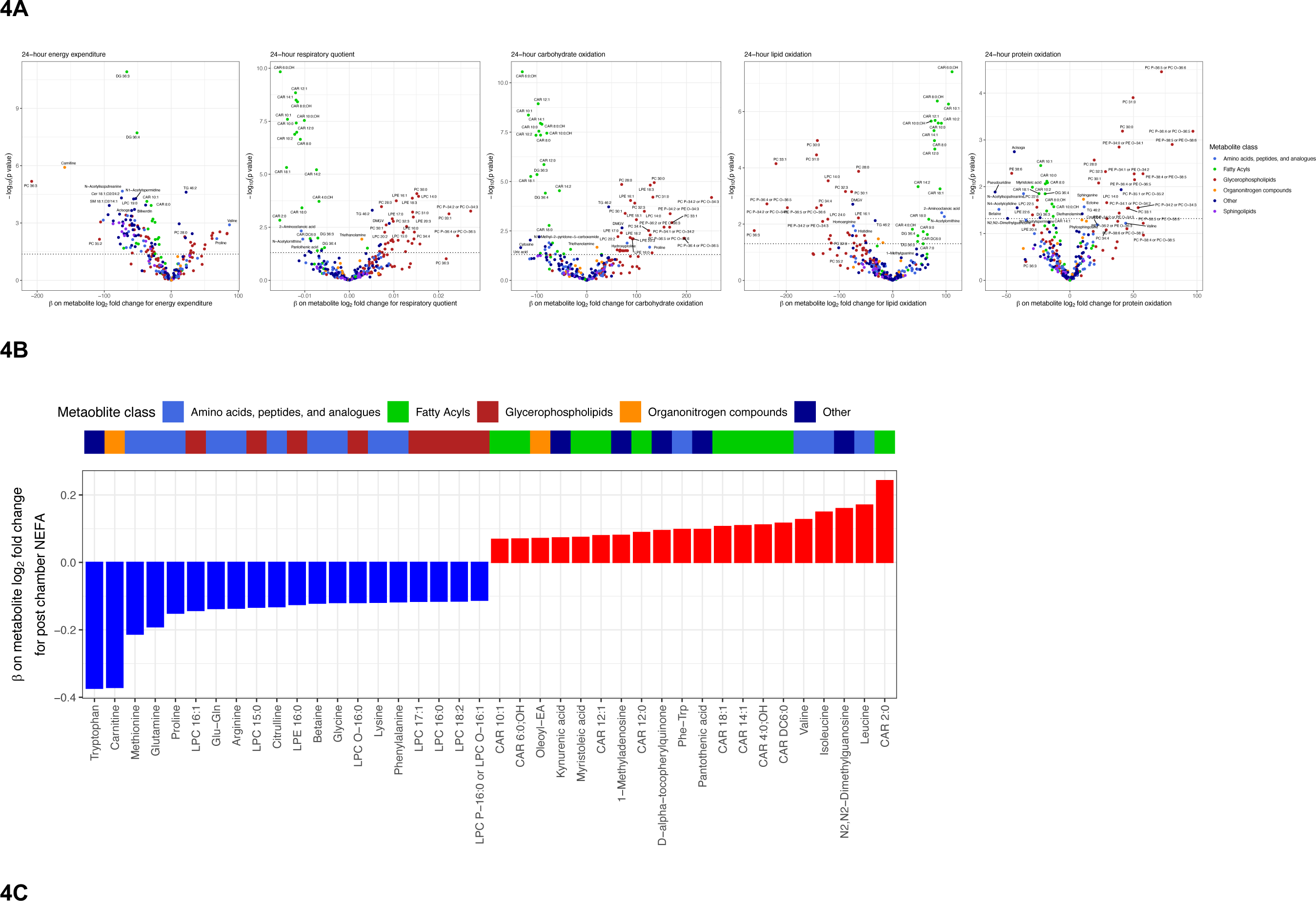
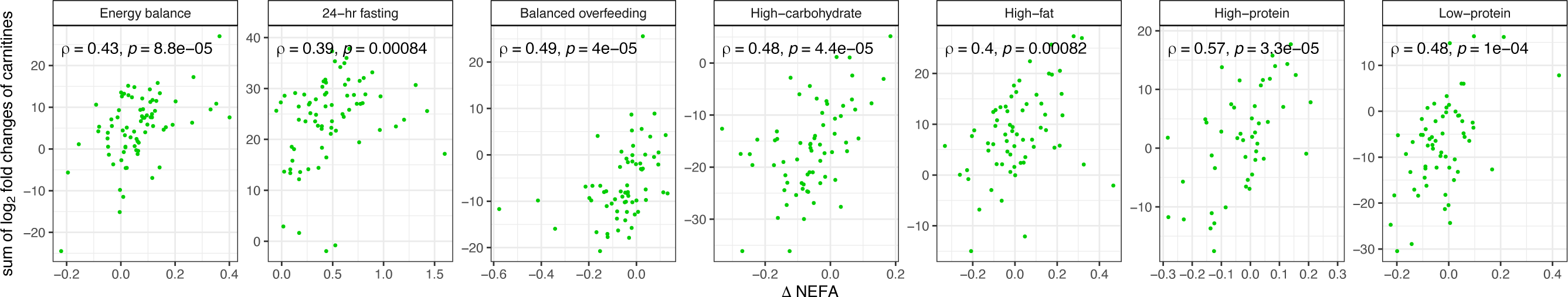
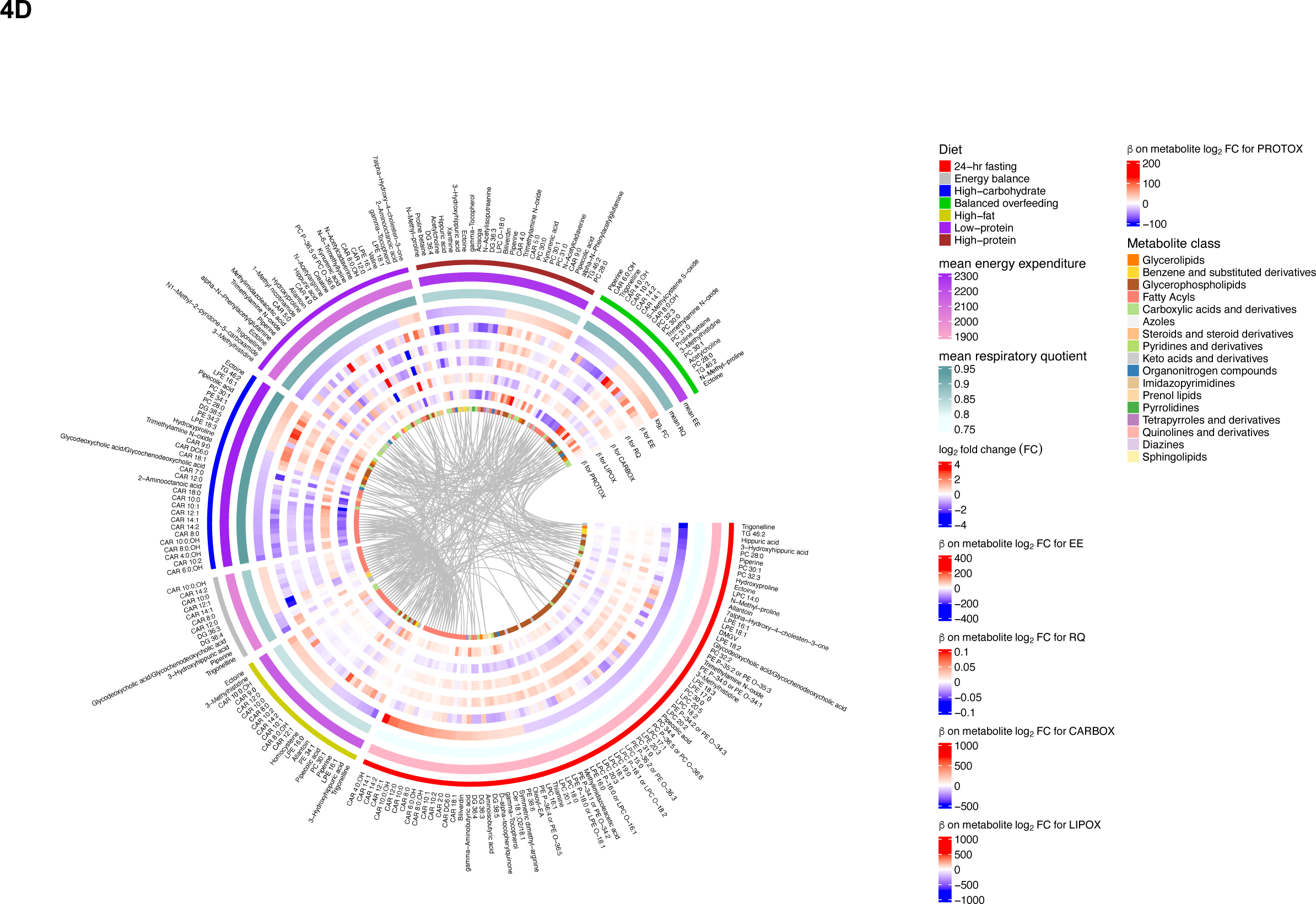
Coordinated metabolomic and physiologic responses to dietary perturbation. Using a linear mixed model for the outcomes of metabolic parameters (e.g., 24-h energy expenditure, 24-h respiratory quotient, etc.), we estimated the effect of log_2_ fold changes in individual metabolites aggregated across diets. Example mixed model: 24-h energy expenditure ∼ log_2_ fold change metabolite + pre-chamber log_2_metabolite level + diet + age + sex + race + BMI + random intercept per participant. **(A)** Volcano plots of the beta coefficients from the log_2_ fold change metabolite variables on respective metabolic parameters. **(B)** Waterfall plot of the beta coefficients on metabolite log_2_ fold change for the outcome of post-chamber non-esterified fatty acids (NEFA). Model: post-chamber NEFA∼ log_2_ fold change metabolite + pre-chamber log_2_ metabolite level + pre-chamber NEFA + diet + age + sex + race + BMI + random intercept per participant. **(C)** Correlation between the sum of log_2_ fold changes in carnitines within participants correlated against the change in NEFA. **(D)** Summary visualization of dietary chamber related changes in metabolites and relations with changes in global measures of metabolism (e.g., energy expenditure, respiratory quotient). For visualization, we present metabolites that with an absolute (mean log_2_ fold change)>0.5 in <u>any</u> diet with an FDR<5%. EE = 24-h energy expenditure (kcal/day), RQ = 24-h respiratory quotient, FC = fold change, CARBOX = 24-h carbohydrate oxidation (kcal/day), LIPOX = 24-h lipid oxidation (kcal/day), PROTOX = 24-h protein oxidation (kcal/day).

Moreover, in examining non-esterified free fatty acid (NEFA) levels pre- and post-chamber as an index of adipose tissue lipid mobilization, we observed a consistent correlation of increased NEFA levels during a chamber with an increase in circulating acylcarnitines, specifically in chambers of caloric excess (Figure 4B-C; **Extended Data Figure 8; Supplemental Tables 8-9**). While our study did not serially sample the metabolome <u>during</u> chambers, these findings do provide support for a balance between increased lipid availability (NEFAs) and its utilization (carnitine metabolism) and liberation (PE and PC metabolism).

Additionally, we observed a positive association between increase in D-alpha-tocopherylquinone and increased NEFA, consistent with its role as a vitamin E catabolite that serves as a carnitine-dependent cofactor for mitochondrial fatty acid desaturases^25^. The relation of NEFA to changes in other species (including amino acids) was complicated and less uniform across diets, likely owing to macronutrient context-specific interactions between intake and metabolism. Unlike 24-h RQ, 24-h EE did not map as consistently to the changes in the assayed metabolome with diet (Figure 4A). Figure 4D represents a full summary of our result, highlighting these shared relations in physiologic-metabolic phenotype across diets.

## Discussion

To our knowledge, this study represents the first application of metabolite profiling to an exquisitely controlled experiment in human metabolism in which individuals were exposed to varied composition of macronutrient overfeeding and fasting in a respiratory chamber. Overall, the results suggest a coordinated structure of intra-individual substrate preferences across diets, which may even be captured in fasting measures. The metabolome is clearly responsive to nutritional interventions^10, 26–28^, but most extant studies have been conducted in less strictly monitored settings than the current report, without phenotyping of individual physiologic responses. In a 4-week study with weight balance and 3 different diets (low fat, low glycemic index, very-low carbohydrate), Esko and colleagues observed a broad change in the metabolome across diets (≈50% of their assayed metabolome)^26^. While the precise dietary exposures were different in nature and duration from our study, some metabolites were shared, including carnitines, select amino acids, ketones, and microbially products. Moreover, while we did not have access to long-term outcomes in the present report, acute metabolic responses to select substrates (e.g., oral glucose^11, 29–31^ or mixed meal tolerance testing^32^) may map to longer-term cardiometabolic outcomes^33^. Indeed, in the present report, we observed dynamic changes in metabolites (glycine^34^, glutamine^35^, DMGV^36–38^) linked to cardiometabolic states in select dietary interventions (e.g., high-fat, fasting, high-protein), opening the potential for individualizing metabolic risk mitigation strategies by precision metabolic response to nutrition.

We acknowledge several key limitations to the study, including modest sample size, limited number of metabolites measured, pre-specification of macronutrient content and off-protocol changes to the specific provided foods (e.g., due to participant preference but without any changes to macronutrient content), carry-forward effects (from inadequate recovery of metabolome between chambers), and measuring post-chamber metabolites nearly approximately 11h after last overfeeding meal. Despite these limitations, this experimental protocol has been used to uncover known functional metabolic mediators of energy expenditure (FGF-21)^39^. While the consistency of signals with fat oxidation and 24-h RQ across diets and evidence of replication in other studies (fasting^10^ and non-fasting^26^) are encouraging, the acknowledged limitations highlight the inherent complexities in studying human nutrition at precision scale, where large studies of prolonged, controlled feeding are logistically, ethically, and financially impractical. In addition, with the advent of high-resolution transcriptomics and proteomics, concomitant tissue sampling during dietary challenge to assess underlying metabolic pathways may serve to further contextualize our findings within the circulating metabolome (e.g., via assessing insulin response, metabolic enzyme expression, mTOR signaling). Overall, these results underscore the importance of detailed physiologic experiments in human nutrition to guide understanding of metabolic mechanisms of dietary response. The upcoming results of large, targeted metabolomic validation efforts of nutrition at population scale (e.g., Nutrition for Precision Health initiative)—contextualized with our results from metabolic chambers—is likely to yield population-level estimates of dietary effects that may reveal how and why individuals respond differently to diet and its implication on metabolic risk.

## Methods

### Study cohort

This study involved a National Institutes of Health (NIH) approved protocol to study short-term metabolic adaptation under macronutrient stress (NIH Protocol #07-DK-N215, PI: Krakoff)^14, 39–41^. The study was approved by the NIH IRB, and all subjects provided written informed consent. Of note, 94 participants had valid data for energy balance respiratory chamber data used to calibrate the 200% caloric excess chambers. Subsequent 24-h intervention diets were randomized. Due to technical issues or participant withdrawal, the number of participants in each dietary chamber varies (Table 1). Participants were required to have a stable weight for 6 months and be otherwise healthy based on medical history and physical examination upon admission to the inpatient NIH Clinical Research Unit. Upon admission, volunteers were placed on a weight maintaining diet (WMD), which was followed prior to and in between the 24-h intervention diets^14^. Participants were weighed daily and calories were adjusted to maintain weight (coefficient of variation of weight over study 0.9±0.6%). After 3 days of the WMD, participants underwent a 75-gram oral glucose tolerance test to exclude individuals with type 2 diabetes (T2D) or impaired glucose regulation (fasting glucose ≥ 100 mg/dl or 2-h glucose ≥140 mg/dl; T2D, fasting glucose ≥126 mg/dl or 2-h glucose 2200 mg/dl). The subjects then underwent a series of 24-h whole-room indirect calorimetry experiments (chambers) to quantify <u>24-h energy expenditure</u> (24-h EE) and <u>24-h respiratory quotient</u> (24-h RQ). A 24-h urine collection was performed to assess protein oxidation rates, and lipid and carbohydrate oxidation were calculated as described^42^. Chambers were performed under several different conditions, including energy balance (calories ingested = calories expended), fasting, and 200% caloric overfeeding diets in random order and with intervening periods of at least 3 days on WMD: (1) standard overfeeding (SOF); (2) a low-protein overfeeding (LPF); (3) high-fat/normal-protein overfeeding (FNP); (4) high-fat/high-protein overfeeding (HPF); (5) high-carbohydrate/normal-protein overfeeding diet (CNP). Caloric excess (200% of individual-specific energy needs) was used to perturb energy balance and to best elicit inter-individual heterogeneity in phenotypes. The total inpatient stay lasted ≈37 days.

### Dietary exposures

Macronutrient composition for each diet is shown in **Supplemental Table 1.** Carbohydrates were a mixture of simple (e.g., soda, candy) and complex (legumes, vegetables, and fruit). Protein source was predominantly of animal origin. Dietary exposures included 24-h fasting and different <u>overfeeding conditions</u> (200% of calories expended during **EB**) and <u>fasting</u> diet macronutrient composition *was* quantified by “The Food Processor” software (ESHA Research, Salem, OR). Residual uneaten food was returned to the metabolic kitchen to calculate actual macronutrient intake during each session. These dietary compositions were chosen specifically to stress the metabolic system not only by providing modern obesogenic diets (e.g. high-carbohydrate or high-fat), but also based on previous data indicating that low-protein (3%) diets amplify inter-individual differences in thermogenesis^43^.

### Metabolic chamber measures

The description of our indirect calorimetry apparatus^44^ and methods^14, 39^ have been published. The first two chamber sessions were used to precisely establish the individual level of energy balance (**EB**), a condition where isocaloric intake matches energy expenditure. Measured 24-h EE during the first eucaloric chamber was used as the energy intake calories for the second chamber to precisely achieve energy balance. This second chamber was used as the EB chamber in this analysis. The order of subsequent chambers was <u>randomized</u> (to limit confounding effects of the order of dietary exposure on metabolism) and spaced by 3-day intervals of WMD and limited physical activity (walking, playing pool, watching TV) to limit “carry-forward” effects from prior a chamber’s diet. Those subjects who could not consume >95% of the food provided by the metabolic kitchen during the chamber sessions were withdrawn from the analysis.

Subjects entered the chamber immediately following breakfast at 7:00 AM (if not a fasting chamber day). Venous blood was drawn before entry into chamber and upon exit into EDTA tubes, with DPP-IV inhibitor and aprotinin, centrifuged for plasma generation and stored at −70°C. Meals in the chamber were provided via a two-way airlock at 11:00 AM, 4:00 PM, and 7:00 PM. Subjects were instructed not to be physically active in the calorimeter to limit the contribution of activity to adaptive thermogenesis. Radar was used to monitor physical activity (denoting % time with motion). The temperature of the chamber was controlled at 24°C, and monthly validation tests (involving propane combustion inside the chamber) verified O_2_ and CO_2_ recovery within 2% based on change in propane weight. Air output from the chamber was sampled every minute and compared to inflow (fresh) air to calculate a subject’s VCO_2_ production and VO_2_ consumption per minute. These measures were utilized to quantify a per-minute **RQ** (= VCO_2_/VO_2_) and the rate of **energy expenditure** [Lusk equation: VO_2_ x 4.686 + (calculated RQ – 0.707) x 0.361/0.293], as described^44, 45^. The per-minute values for energy expenditure and RQ were extrapolated to 24-h (multiplied by 1440). 24-h urinary nitrogen excretion rate was measured while in the chamber to estimate 24-h protein oxidation rate and used to derive the non-protein RQ, which was used to calculate 24-h carbohydrate (CARBOX) and lipid (LIPOX) oxidation rates^42^ as a secondary measure of substrate oxidation preference. We have demonstrated excellent reproducibility of metabolic measures previously^41^.

### Metabolite profiling

Measurement of 322 metabolites, including amino acids, acylcarnitines, and other cationic polar metabolites, were made using liquid chromatography– tandem mass spectroscopy (LC-MS) as described previously^45^. We observed an excellent coefficient of variation in pooled QC samples across metabolites using raw, not log-transformed, data (median 4.7%, 25^th^-75^th^ percentile, 3.4%-7.6%). We reviewed distributions of the pre-chamber metabolite levels for each dietary chamber and excluded measurements that were >5 standard deviations within a diet. We excluded metabolites with any degree of missingness from our analysis (including those missing due to outlier removal), leaving 263 metabolites for analysis. We log_2_ transformed metabolite levels prior to use in models for model interpretation to be centered around log_2_ fold changes in metabolite levels.

### Statistical analysis

While our sample size is large for studies of this degree of precision phenotyping in 24-h metabolic chambers, we were sensitive to dimensionality and overfitting concerns given the number of metabolites and chambers. In this regard, we performed two approaches to test the effect of each dietary chamber on changes in metabolite levels: a serial pre-vs. post-chamber metabolite comparison (*t*-test) and a more complex mixed model approach including interaction terms to model random chamber order and account for cross-over effects. For *t*-tests, A false discovery of 5% (Benjamini-Hochberg) was imposed across all tested metabolites within a dietary chamber. To estimate the effect of each dietary chamber on the log_2_ fold change in metabolite (in relation to the energy balance chamber), we constructed linear mixed models of the form: post-chamber metabolite log_2_ level ∼ pre-chamber log_2_ metabolite level + diet + chamber order + random effect per participant. Chamber order refers to the order the participant entered the dietary chamber (e.g., energy balance is 1 for all participants as this was the first chamber entered, the remaining chambers are in random orders across participants). The diet variable was structured with energy balance diet as the referent.

To test the relations of changes in metabolites with physiologic measures including 24-h energy expenditure, 24-h respiratory quotient, and oxidation sub-types, we used linear mixed models. This approach combines data from all dietary chambers into one model with repeated measures. As an example, the model for 24-h energy expenditure was of the following form: 24-h energy expenditure ∼ log_2_ fold change metabolite + pre-chamber metabolite log_2_ level + diet + age + sex + race + BMI + random effect per participant. We compared these models to linear models, stratified by diet of the following form: 24-h energy expenditure ∼ log_2_ fold change metabolite + pre-chamber log_2_ metabolite level + age + sex + race + BMI, where each model was restricted to data from a single dietary chamber. To test the relations of changes in metabolites with changes in non-esterified free fatty acids (NEFA), we created analogous sets of mixed and linear models, for the outcome of post-chamber NEFA with adjustment for pre-chamber NEFA, pre-chamber log_2_ metabolite level, age, sex, race, and BMI (the mixed model again had a random effect per participant).

## Data availability

The metabolic and phenotypic data from this study are available via controlled access at NIH (contact: Dr. Jonathan Krakoff) due to participant confidentiality and inclusion of protected groups. Requests for data should be made to Dr. Jonathan Krakoff directly, with subsequent approval by NIH. Code utilized in these analyses are available at https://github.com/asperry125/MetFlex.

---

**Extended Data Figure 1:**
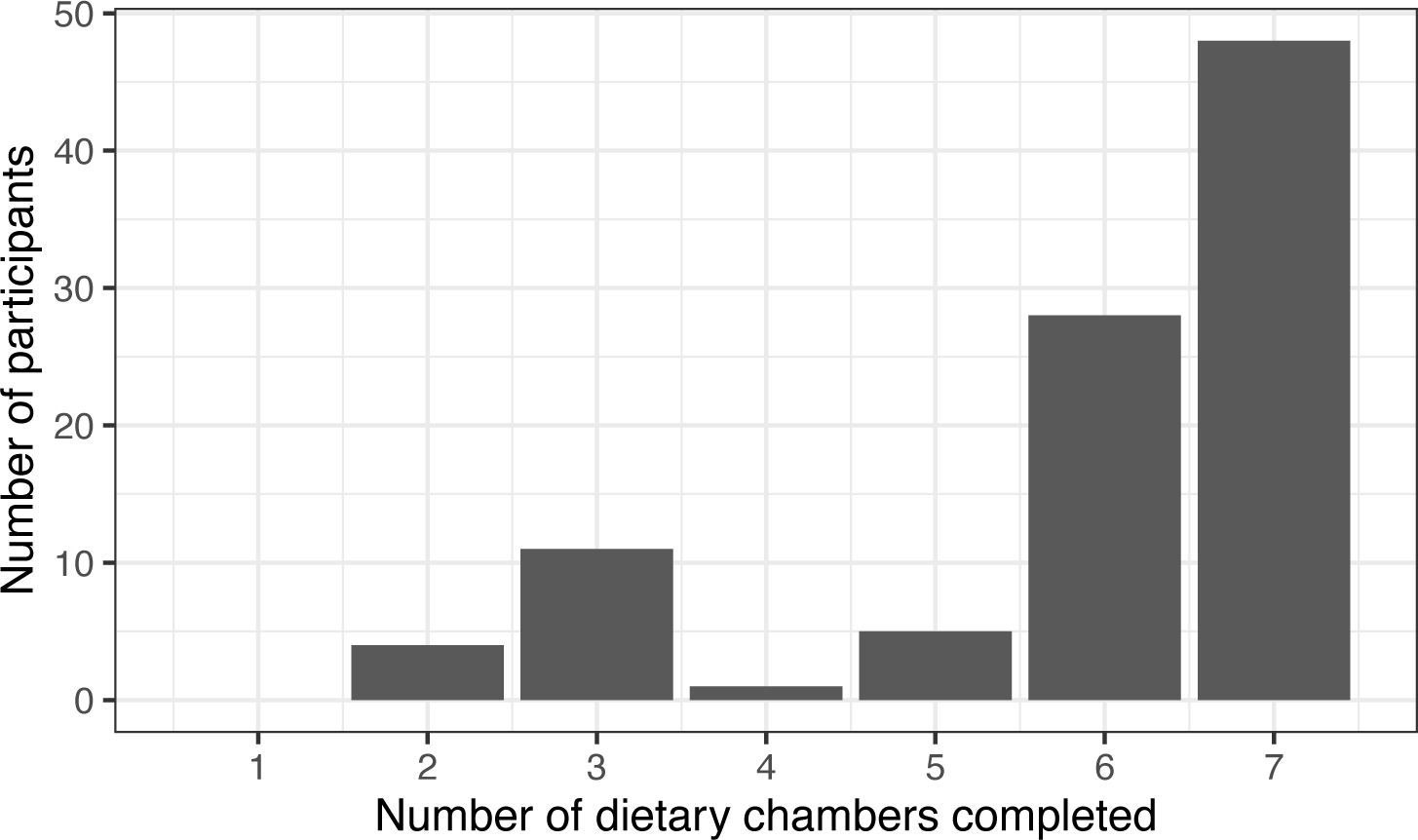
Number of dietary chambers completed by study participants.

**Extended Data Figure 2:**
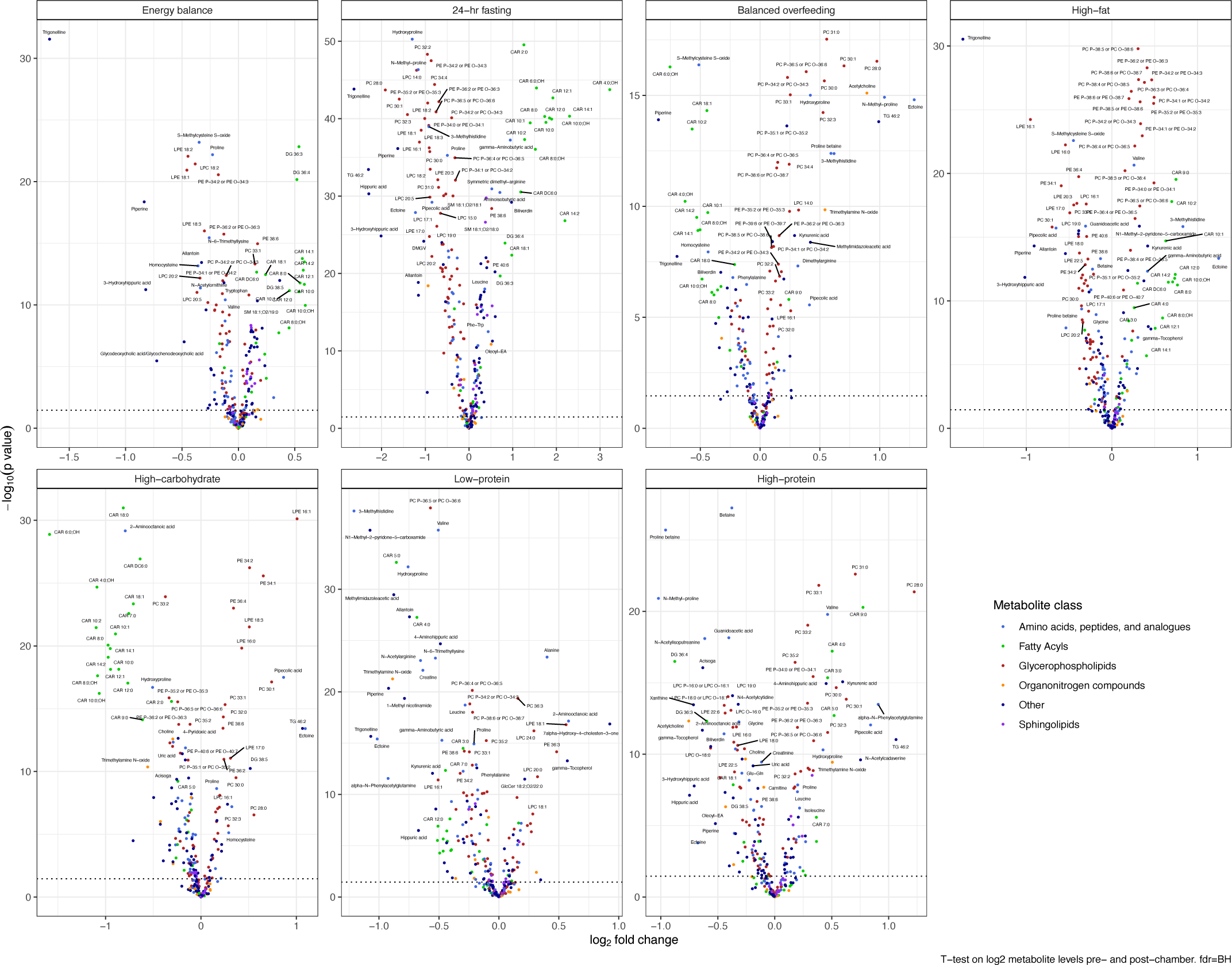
Full results from *t*-tests across all 7 dietary chambers.

**Extended Data Figure 3:**
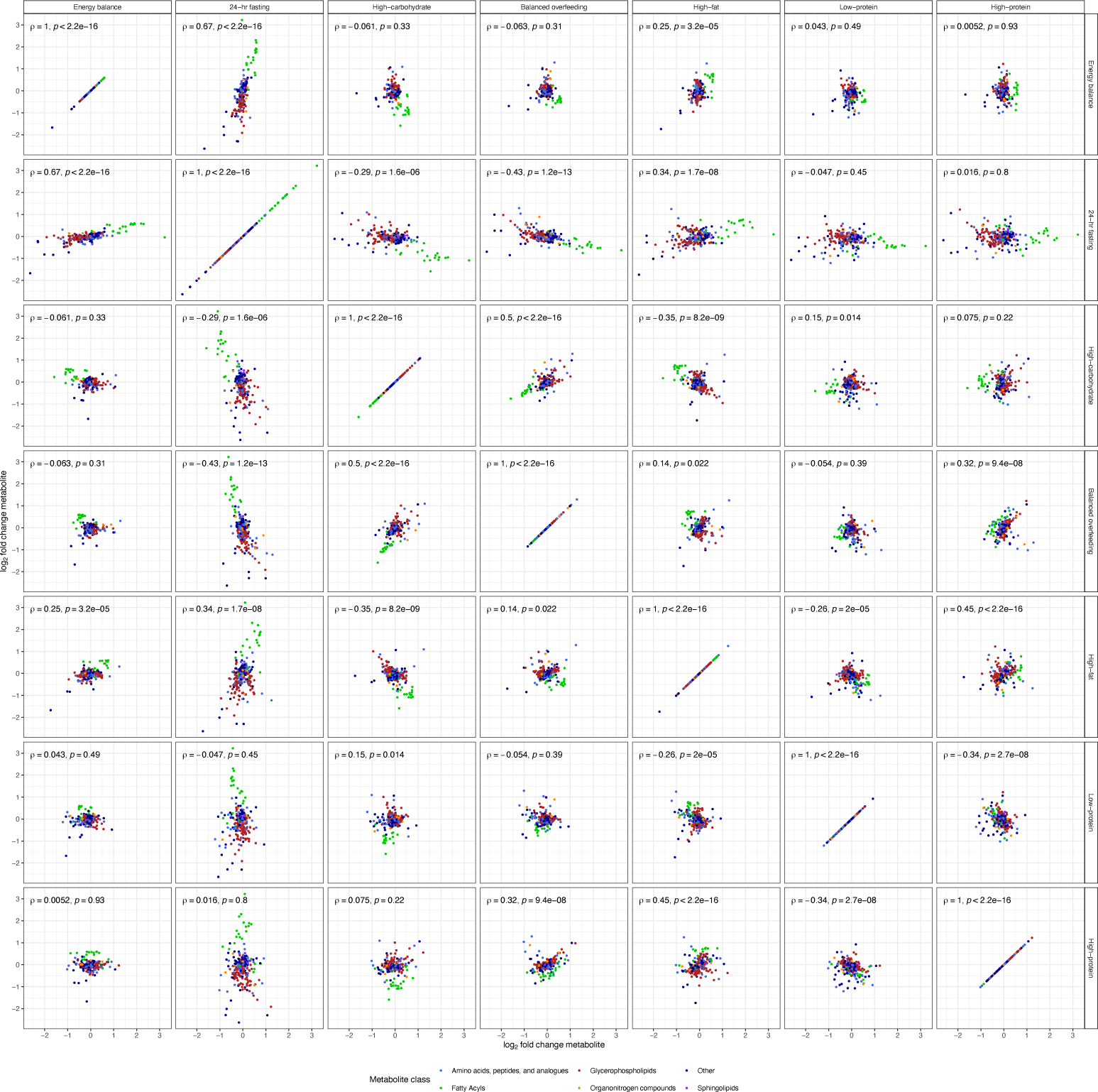
Correlation of mean log_2_ fold changes across all dietary chambers. Spearman correlation plots comparing all diets against each other. This presents the full data which is summarized in **Figure 3D**.

**Extended Data Figure 4.**
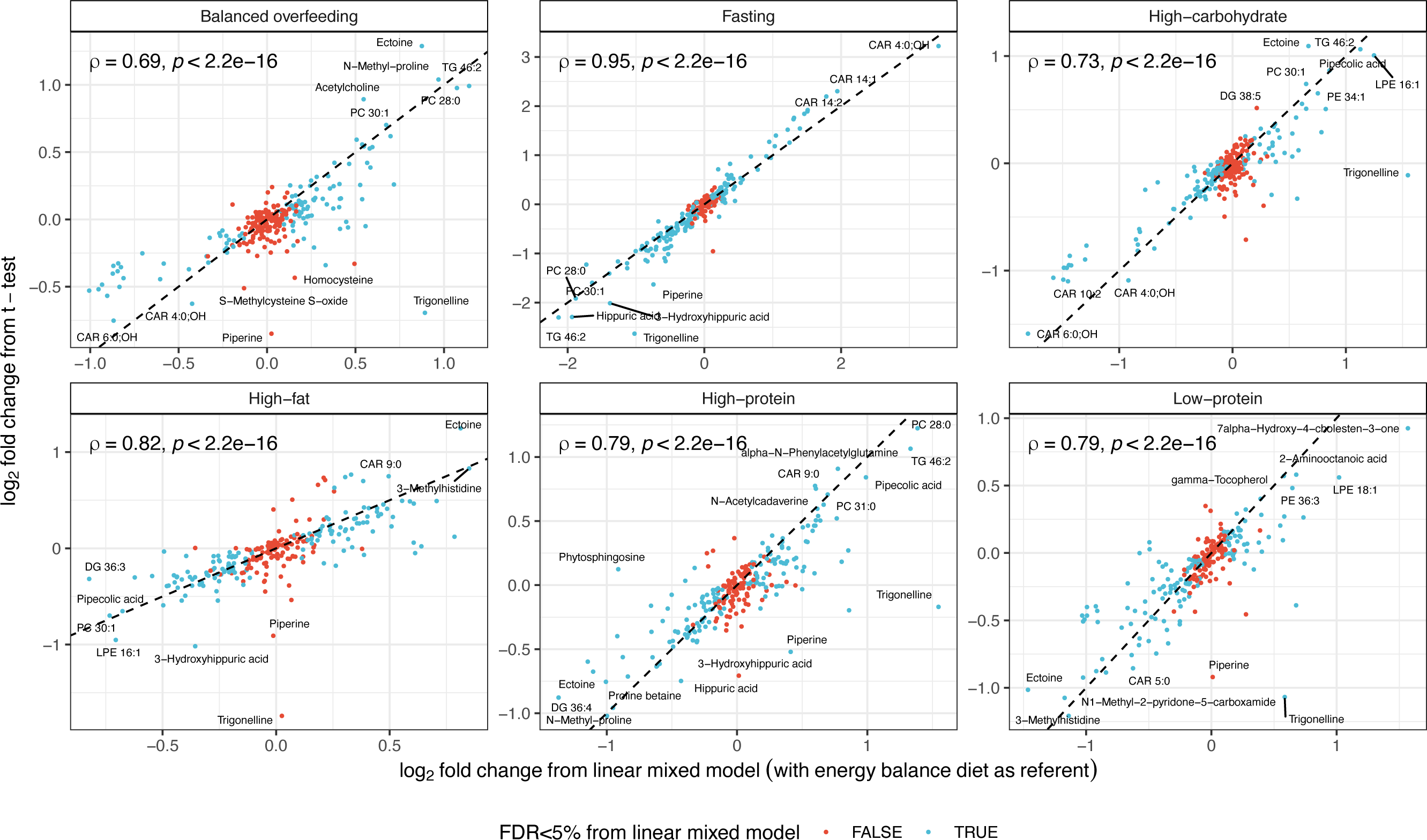
Comparison of estimated log_2_ fold change in metabolite from *t*-tests and a linear mixed model. Paired *t*-tests demonstrated numerous changes to circulating metabolites for each dietary chamber (**Figure 3A-B**). In order to estimate the effect of each dietary chamber on the log_2_ fold change in metabolite, we constructed linear mixed models as follows: post-chamber log_2_ metabolite level ∼ pre-chamber log_2_ metabolite level + diet + chamber order + random effect per participant. Chamber order refers to the order the participant entered the dietary chamber (e.g., energy balance is 1 for all participants as this was the first chamber entered, the remaining chambers are in random orders across participants). The diet variable was structured with energy balance diet as the referent. The figure presents the estimated effect of the dietary chamber on the log_2_ fold change of each metabolite, compared to the energy balance chamber (x-axis). These effects are compared against what was obtained via paired *t*-tests (y-axis).

**Extended Data Figure 5.**
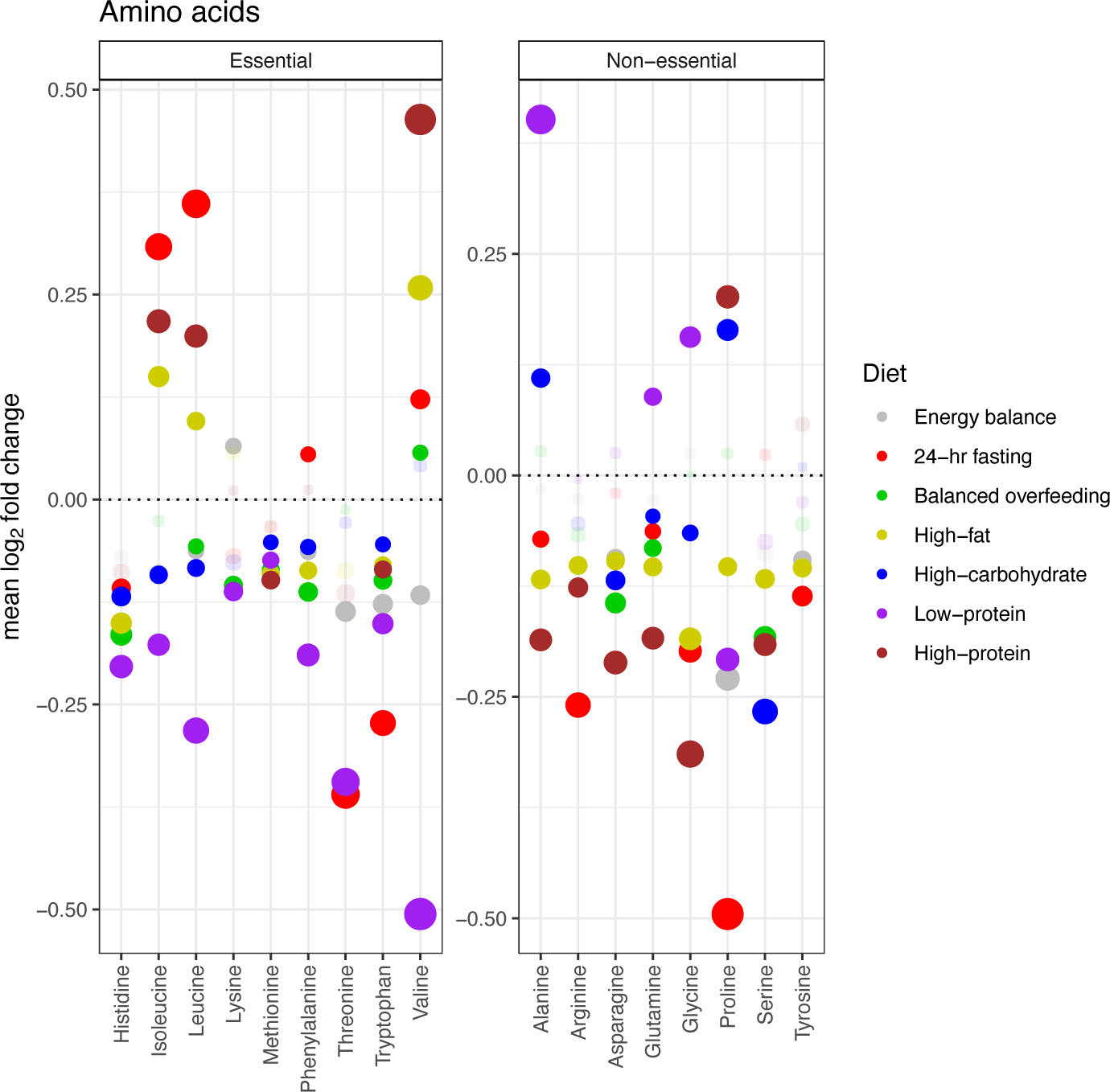
Changes in essential v. non-essential amino acids. Comparison of the mean log_2_ fold change of essential and non-essential amino acids (which were quantified in our study) across all 7 dietary chambers. Colors represent the dietary chamber, size is proportional to mean log_2_ fold change, and points are faded if the mean log_2_ fold change for the diet-metabolite combination did not reach an FDR<5%.

**Extended Data Figure 6A:**
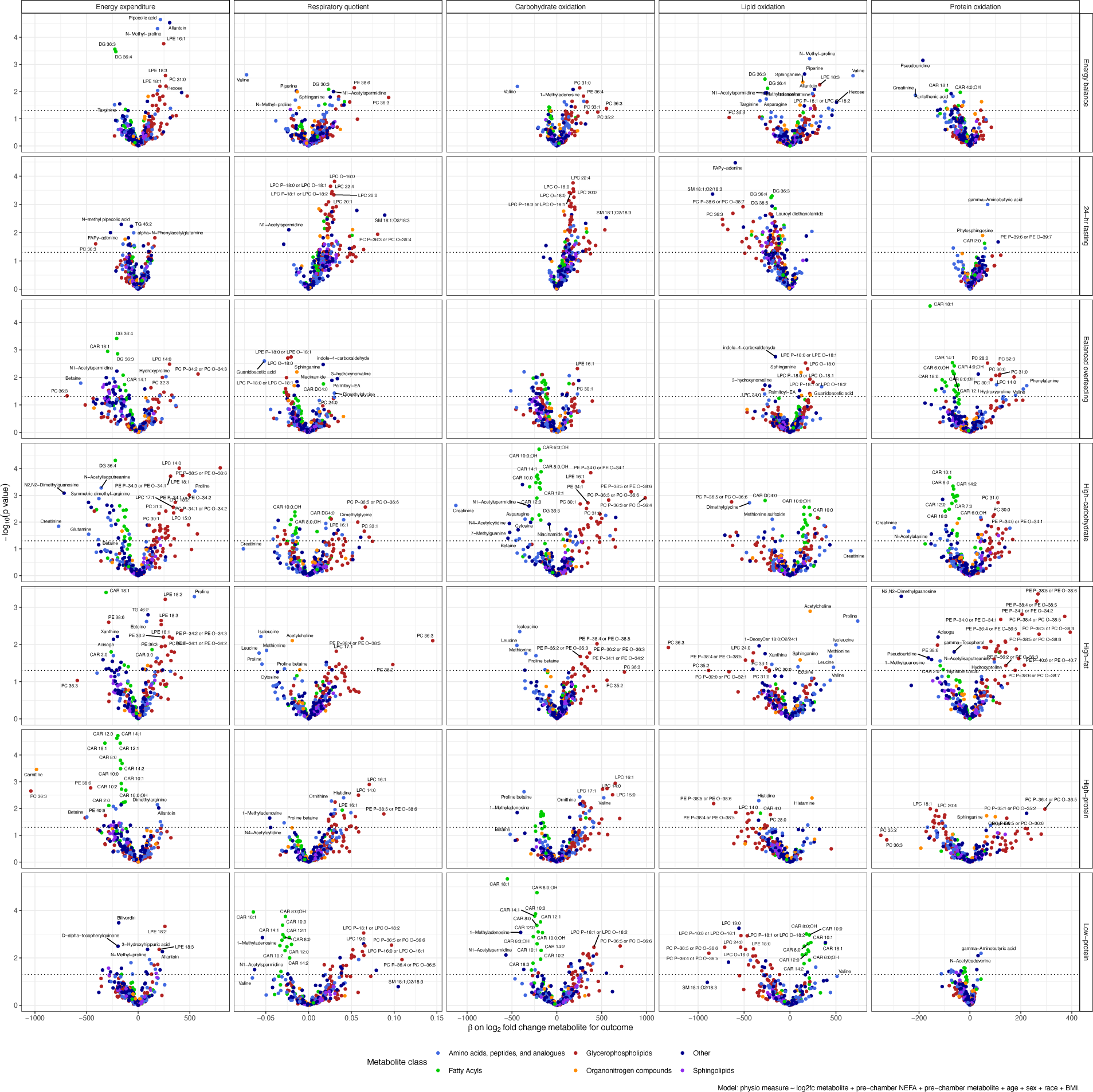
Relationship between log_2_ fold change metabolite and physiologic measures by diet. Volcano plots reporting the beta coefficient on log_2_ fold change metabolite as a predictor of physiologic measures, such as energy expenditure and respiratory quotient. Example model: 24-h energy expenditure ∼ log_2_ fold change metabolite + pre-chamber metabolite + age + sex + race + BMI.

**Extended Data Figure 6B:**
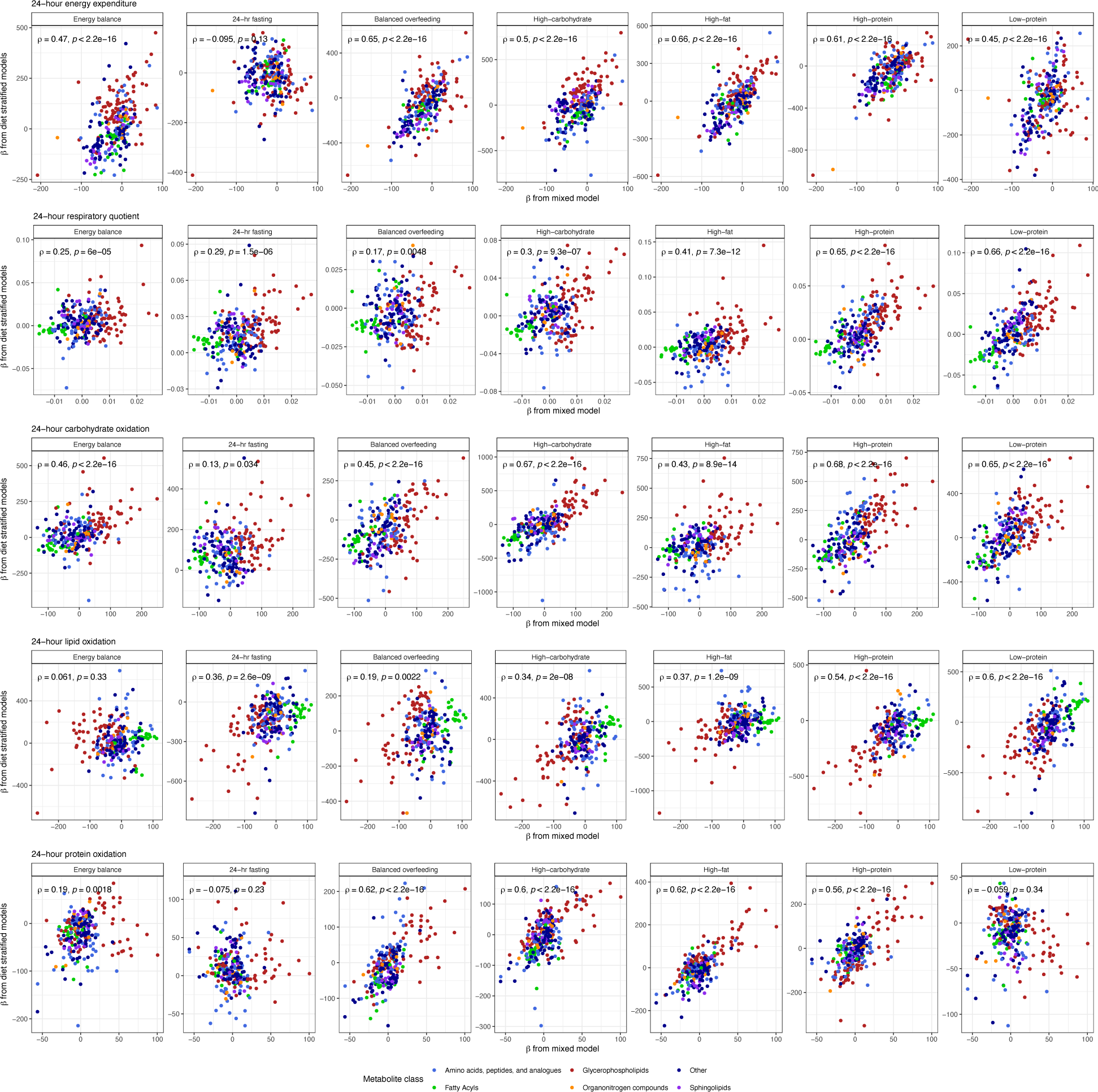
Comparison of mixed model with diet stratified linear models. Beta coefficients on the x-axes come from linear mixed models of the structure outcome (e.g., energy expenditure) ∼ log_2_ fold change metabolite + pre-chamber metabolite + diet + age + sex + race + BMI + random effect per participant. Beta coefficients on the y-axes come from diet stratified linear models of the structure outcome ∼ log_2_ fold change metabolite + pre chamber metabolite + age + sex + race + BMI.

**Extended Data Figure 7:**
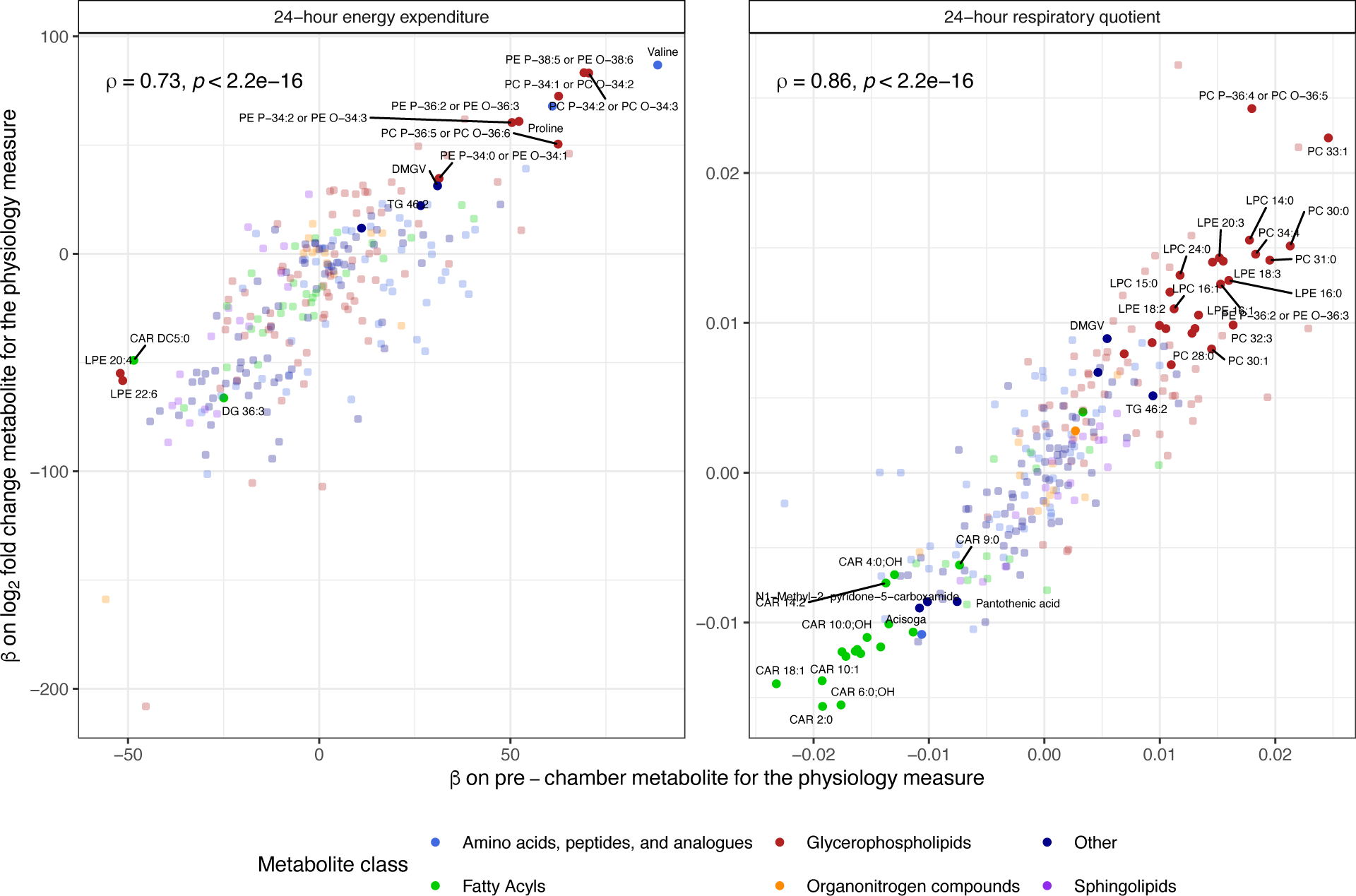
Comparison of effects of pre-chamber metabolite levels and log_2_ fold change metabolite on physiologic measures. Scatterplots comparing beta coefficients from the pre-chamber metabolite variable and the log_2_ fold change metabolite variable for linear mixed models. Points are faded if neither beta coefficient had an FDR<5%.

**Extended Data Figure 8:**
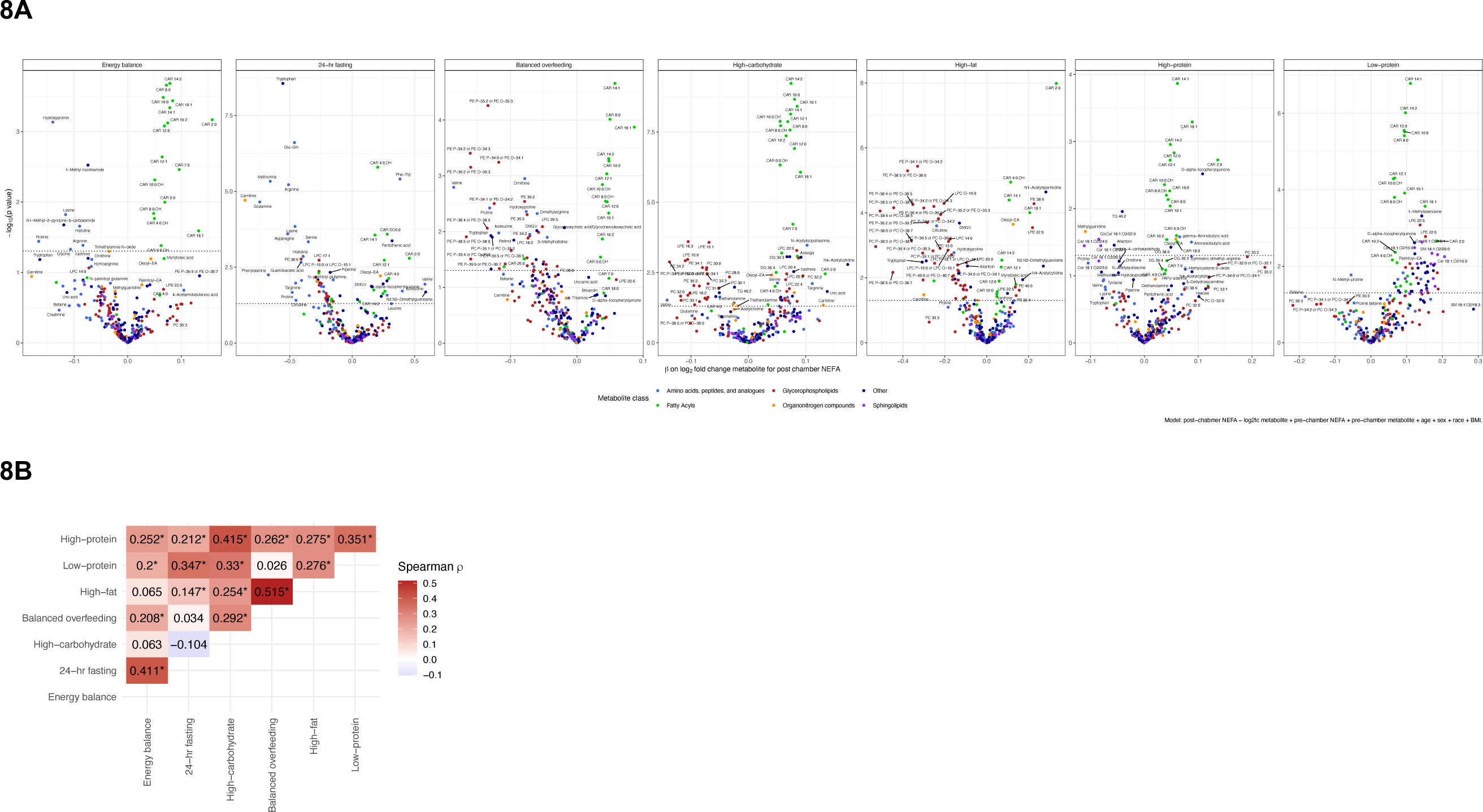
Relations between log_2_ fold change metabolite and change in non-esterified free fatty acids (NEFA) in diet-stratified models. **(A)** Diet stratified models for NEFA with beta coefficients on the log_2_ fold change metabolite. **(B)** Spearman correlation heatmap of the beta coefficient on log_2_ fold change metabolite on the outcome of post-chamber NEFA (adjusted for pre-chamber NEFA and pre chamber metabolite and age/sex/race/BMI) across dietary chambers.

